# Extracorporeal Shockwave Therapy Versus Ultrasound Therapy for Plantar Fasciitis: Systematic Review and Meta-Analysis

**DOI:** 10.1101/2020.09.20.20198168

**Authors:** Zeyana Al-Siyabi, Mohammad Karam, Ethar Al-Hajri, Abdulmalik Alsaif

## Abstract

**Objective:** To compare the outcomes of Extracorporeal Shockwave Therapy (ESWT) versus Ultrasound Therapy (UST) in plantar fasciitis.

**Methods:** A systematic review and meta-analysis were performed. An electronic search identifying studies comparing ESWT and UST for plantar fasciitis was conducted. Primary outcomes were morning and activity pain, functional impairment and the American Orthopaedic Foot and Ankle Society (AOFAS) scale score. Secondary outcomes included fascial thickness, primary efficacy success rate, activity limitations, pain intensity and satisfaction.

**Results:** Seven studies enrolling 369 patients were identified. No significant difference was found between ESWT and UST for functional impairment (Mean Difference [MD]= -2.90, P= 0.22), AOFAS scale score (MD= 35, P= 0.20) and pain in the first steps in the morning (MD= -4.72, P= 0.39). However, there was a significant improvement in pain during activity for the ESWT group (MD= -1.36, P= 0.005). For secondary outcomes, ESWT had improved results in terms of primary efficacy success rate, activity limitations and patient satisfaction. Reduction of planter fascia thickness showed no significant difference. Pain intensity after treatment had varied results amongst included studies.

**Conclusion:** ESWT is superior to UST for plantar fasciitis as it improves pain activity and intensity, primary efficacy success rate and activity limitations.

## Introduction

Plantar fasciopathy or plantar fasciitis is one of the most common foot disorders that occurs in approximately 10% of the population throughout their life^1^. Although it was defined as an inflammatory syndrome, recent studies have emphasised that plantar fasciopathy is more likely to be a degenerative process associated with multi-factorial aetiology^2,3^. Factors are thought to be anatomical (such as pes planus and pes cavus) or biomechanical (such as excessive external rotation and subtalar joint overpronation) or environmental (such as obesity and inappropriate footwear)^4–7^. Plantar fasciitis is diagnosed clinically and MRI imaging is a second-line diagnostic test to confirm diagnosis and rule-out other foot disorders due to its considerable costs^8,9^. Typical presentations are throbbing, burning, or piercing medial heel pain, especially in the first steps in the morning or after prolonged rest period^3^. The pain typically decreases after a few steps but may return with continued weightbearing activities^3^. If untreated, pain may last for months or years^3^. Conservative treatments (such as activity modification, oral analgesics, ice massage, stretching exercises, orthotics and corticosteroid injections) can help with the disabling pain. Patients with chronic plantar fasciitis can consider other treatment options, including extracorporeal shockwave therapy (ESWT), ultrasound therapy (UST), low-level laser therapy (LLLT) or surgical plantar fasciotomy^10,11^.

ESWT comprises focused pulses of high-pressure sound waves to bombard damaged tissues aiming to minimise pain and symptoms associated with plantar fasciitis. They were initially used for medical purposes in the management of renal calculi by lithotripsy. Subsequently, shockwaves were utilised in the treatment of ununited fractures^12,13^. Years later, they became popular in Germany for certain musculoskeletal complains, including calcifying tendonitis epicondylitis and plantar fasciitis^14^. ESWT has been used as an alternative to surgery for patients with long-term, recalcitrant planter heel pain. The mechanism of the action of shock waves on soft-tissue conditions remain speculative. Experts propose the pulses to bombard the central nervous system by causing alterations in the permeability of cell membranes inhibiting the transmission of painful stimuli resulting in pain relief, while others contend that they stimulate the healing cascades by essentially re-injuring the tissues^15,16^.

Therapeutic ultrasound (US) is one of the physiotherapeutic modalities commonly used in the management of soft tissue disorders such as planter fasciitis. US is a high-frequency wave that produces thermal or non-thermal effects depending on the frequency, intensity, duration of pulses and injury type^17^. It was reported that ultrasound has advantages on the healing of soft tissue^18,19^. It has a base unit for generating electrical signals that transmit through biologic tissues causing a raise in tissue temperature and metabolism and thus enhancing blood circulation^20^. Ultrasonic energy has also been purported to affect the chemical activity of tissues by increasing the permeability of the cell membranes and regulating the molecular structures and protein production, all possibly resulting in the promotion of tissue recovery and shorter healing process^19^. Nonetheless, there is a lack of high-quality scientific evidence to support the practical use of UST in the management of musculoskeletal conditions.

There are currently no systematic reviews or meta-analyses that solely compare the use of ESWT against UST for plantar fasciitis treatment although they have been reported in several recent randomised controlled trials (RCTs) as well as non-randomised cohort studies^21-27^. Therefore, it is imperative to conduct the first review within the literature regarding this topic.

## Methods

A systematic review and meta-analysis were conducted as per the Preferred Reporting Items for Systematic Reviews and Meta-Analyses (PRISMA) guidelines^28^.

### Eligibility criteria

All RCTs and observational studies comparing ESWT with UST for plantar fasciitis treatment were included. ESWT was the intervention group of interest and UST was the comparator. All patients were included irrespective of age, gender or co-morbidity if they belonged to either a study or control group. Case reports and cohort studies where no comparison was conducted were excluded from the review process.

### Primary Outcomes

The primary outcomes are pain in the morning and during activity, functional impairment and the American Orthopaedic Foot and Ankle Society (AOFAS) scale score. Pain was reported using a visual analogue scale (VAS) during the morning pain when taking the first steps and during activity like exercise or walking ^23, 24, 26^. Using the self-administered questionnaire “University of Peloponnese Pain, Functionality and Quality of Life Questionnaire”, functional impairment was evaluated on a five-point Likert scale, before treatment, immediately after and at 4-week follow-up^27^. The AOFAS scale was used to measure foot functionality and range of motion^24^.

### Secondary Outcomes

The secondary outcomes included are fascial thickness before and after treatment, primary efficacy success rate, activity limitations, pain intensity and patient satisfaction.

### Literature search strategy

Two authors independently searched the following electronic databases: MEDLINE, EMCARE, EMBASE, CINAHL, and the Cochrane Central Register of Controlled Trials (CENTRAL). The last search was run on the 29^th^ of March 2020. Thesaurus headings, search operators and limits in each of the above databases were adapted accordingly. In addition, World Health Organization International Clinical Trials Registry (http://apps.who.int/trialsearch/), ClinicalTrials.gov (http://clinical-trials.gov/), and ISRCTN Register (http://www.isrctn.com/) were searched for details of ongoing and unpublished studies. No language restrictions were applied in our search strategies. The search terminologies included “Extracorporeal Shockwave Therapy”, “Ultrasound Therapy”, “Plantar Fasciitis” and “Plantar fasciosis”. The bibliographic lists of relevant articles were also reviewed.

### Selection of Studies

The title and abstract of articles identified from the literature searches were assessed independently by two authors. The full texts of relevant reports were retrieved and those articles that met the eligibility criteria of our review were selected. Any discrepancies in study selection were resolved by discussion between the authors.

### Data Extraction and Management

An electronic data extraction spreadsheet was created in line with Cochrane’s data collection form for intervention reviews. The spreadsheet was pilot-tested in randomly selected articles and adjusted accordingly. Our data extraction spreadsheet included study-related data (first author, year of publication, country of origin of the corresponding author, journal in which the study was published, study design, study size, clinical condition of the study participants, type of intervention, and comparison), baseline demographics of the included populations (age and gender) and primary and secondary outcome data. Two authors cooperatively collected and recorded the results and any disagreements were solved via discussion.

### Data synthesis

Data synthesis was conducted using the Review Manager 5.3 software. The extracted data was entered into Review Manager by two independent authors. The analysis involved used was based on fixed and random effects modelling. The results were reported in forest plots with 95% Confidence Intervals (CIs).

For continuous outcomes, the Mean Difference (MD) was calculated between the 2 groups. A positive MD for the pain, functional impairment and AOFAS scale score would favour the ESWT group, a negative MD would favour the US therapy group and a MD of 0 would favour neither groups.

### Assessment of Heterogeneity

Heterogeneity among the studies was assessed using the Cochran Q test (χ2). Inconsistency was quantified by calculating I^2^ and interpreted using the following guide: 0% to 25% may represent low heterogeneity, 25% to 75% may represent moderate heterogeneity, and 75% to 100% may represent high heterogeneity.

## Results

### Literature search results

The search strategy retrieved 63 studies and after a thorough screening of the retrieved articles, seven studies in total were identified which met the eligibility criteria (Figure 1).

**Figure 1:**
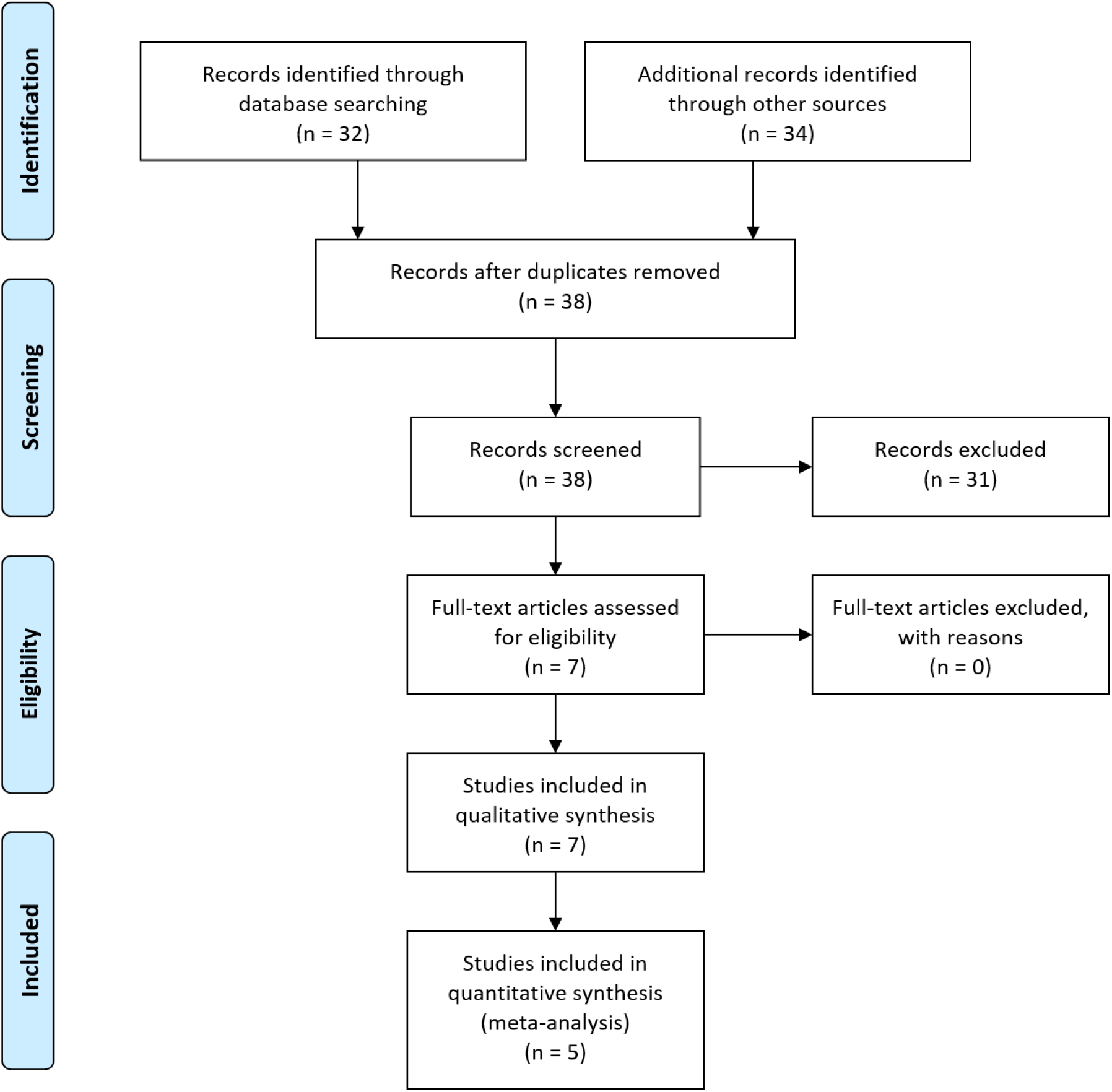
Prisma Flow Diagram. The PRISMA diagram details the search and selection processes applied during the overview. PRISMA, Preferred Reporting Items for Systematic Reviews and Meta-Analyses.

### Description of Studies

Baseline characteristics of the included studies are summarised in Table 1.

**Table 1.** Baseline Characteristics of the Included Studies.

| Study | Year | Country | Journal | Study Design | Total Study Sample | Interventions Compared |
| --- | --- | --- | --- | --- | --- | --- |
| Cheing et al. <sup>21</sup> | 2007 | China | Shock Waves | Cohort Study | 37 | ESWT vs. UST vs. no treatment |
| Greve et al. <sup>22</sup> | 2009 | Brazil | Clinics | RCT | 32 | rESWT vs. UST |
| Konjen et al. <sup>23</sup> | 2015 | Thailand | Journal of the Medical Association of Thailand | RCT | 30 | rESWT vs. UST |
| Ulusoy et al. <sup>24</sup> | 2017 | Turkey | The Journal of Foot & Ankle Surgery | RCT | 60 | ESWT vs. UST vs. LLLT |
| Akinoğlu et al. <sup>25</sup> | 2017 | Turkey | Pain Medicine | RCT | 54 | rESWT + exercise vs. UST + exercise vs. exercise |
| Akinoğlu <sup>26</sup> | 2018 | Turkey | Journal of Exercise Rehabilitation | RCT | 54 | rESWT + exercise vs. UST + exercise vs. exercise |
| Dedes et al. <sup>27</sup> | 2019 | Greece | Acta Informatica Medica | Cohort Study | 159 | rESWT vs. UST vs. conventional therapy |

#### Cheing et al. ^21^

Cheing et al. conducted a dual-centre prospective cohort study that included 37 participants with chronic plantar fasciitis. Participants were allocated into one of three groups, which received ESWT, US or no treatment (control). Only one of the two participating clinics was equipped with the ESWT machine and hence patients attending this clinic were allocated to the ESWT group (12 patients). Patients in the second clinic were randomly assigned to either the US (15 patients) or to the control group (10 patients) by drawing lots.

#### Greve et al. ^22^

Greve et al. conducted a single centre randomised, prospective and comparative clinical study that included 32 patients with chronic plantar fasciitis. Participants were divided into two groups in accordance with randomly drawn numbers: 16 in rESWT and 16 in UST.

#### Konjen et al. ^23^

Konjen et al. conducted a single centre prospective randomised clinical trial that included 30 patients with chronic plantar fasciitis. A computerised random number generator was used to conduct block randomisation into two groups: 15 patients in radial ESWT (rESWT) and 15 patients in UST group.

#### Ulusoy et al. ^24^

Ulusoy et al. conducted a single centre prospective randomised clinical trial that included 60 patients with chronic recalcitrant plantar fasciitis. Using the stratified block randomization method according to gender and body mass index, participants were randomised into 3 treatment groups. 20 patients in the ESWT group, 20 in the UST and 20 in the LLLT.

#### Akınoğlu et al. (2017) ^25^

Akınoğlu et al. (2017) conducted a single centre prospective randomised controlled trial that included 54 patients with chronic plantar fasciitis attending Physical Medicine and Rehabilitation clinic. Sealed envelope method was used for randomisation of study sample into three groups: 24 patients in the rESWT and exercise group, 26 patients in the UST and exercise group and 28 patients in the exercise group.

#### Akınoğlu (2018) ^26^

Akınoğlu (2018) conducted a single centre prospective randomised controlled trial that included 54 patients with chronic plantar fasciitis attending Physical Medicine and Rehabilitation clinic. This is the same study group as Akınoğlu et al. (2017) ^25^; however, the more recent study^26^ reported different outcomes. Sealed envelope method was used for randomisation of study sample into three groups: 24 patients in the rESWT and exercise group, 26 patients in the UST and exercise group and 28 patients in the exercise group.

#### Dedes et al. ^27^

Dedes et al. performed a single centre prospective cohort study that included 156 patients with chronic plantar fasciitis attending an orthopaedic clinic. The study period was from February 2015 to August 2017. The study groups included: 88 patients for rESWT group, 56 patients for UST group and 15 patients for the control group.

### Primary Outcomes

#### Morning and Activity Pain

In Figure 2, morning pain was reported in Konjen et al. ^23^, Ulusoy et al.^24^ and Akınoğlu (2018)^26^ enrolling 73 patients. There was no statistically significant difference seen in the mean difference analyses showing a lower level of pain in the morning for the ESWT group (MD = -4.72, CI = -15.59 to 6.15, P = 0.39). A high level of heterogeneity was found amongst the studies (I^2^ = 100%, P < 0.00001).

**Figure 2:**
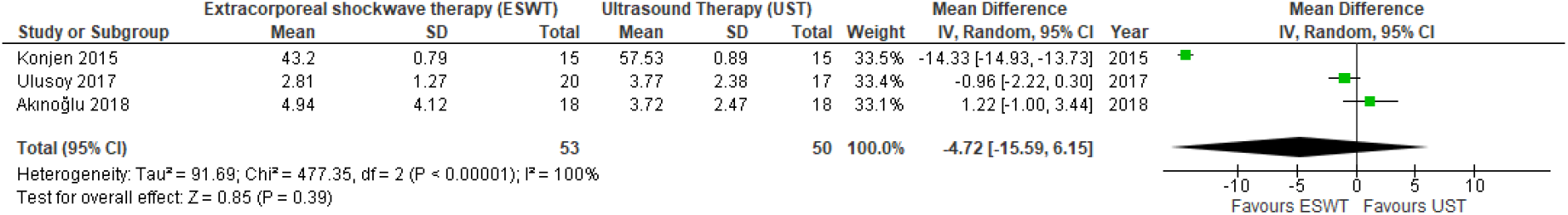
Forest Plot of ESWT vs. UST – Morning Pain. Quantitative analysis showing the mean difference in pain in the morning reported by Konjen et al. (2015), Ulusoy et al. (2017) and Akınoğlu (2018). ESWT and UST stand for extracorporeal shockwave therapy and ultrasound therapy, respectively.

In Figure 3, activity pain was reported in Ulusoy et al.^24^ and Akınoğlu (2018)^26^ enrolling 73 patients. There was a statistically significant difference seen in the mean difference analyses showing a lower level of pain during activity for the ESWT group (MD = -1.36, CI = -2.30 to -0.41, P = 0.005). A medium level of heterogeneity was found amongst the studies (I^2^ = 73%, P = 0.06).

**Figure 3:**
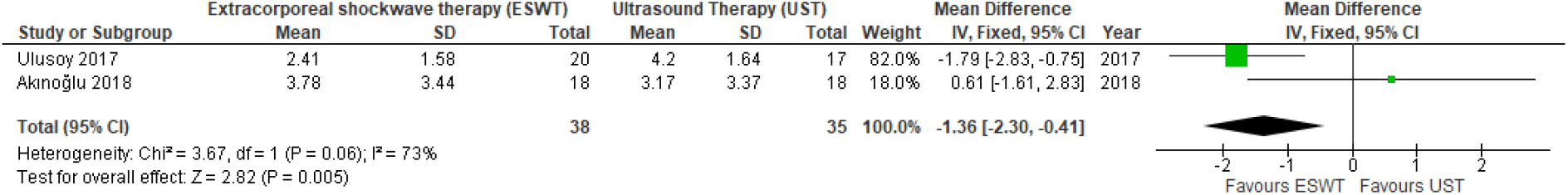
Forest Plot of ESWT vs. UST – Activity Pain. Quantitative analysis showing the mean difference in pain during activity reported by Ulusoy et al. (2017) and Akınoğlu (2018). ESWT and UST stand for extracorporeal shockwave therapy and ultrasound therapy, respectively.

#### Functional Impairment

In Figure 4, functional impairment was reported by Konjen et al.^23^, Akınoğlu et al. (2017) ^25^ and Dedes et al.^27^ enrolling 106 patients. There was no significant statistical difference seen in the standard mean difference analyses showing a higher functional impairment in the ESWT group compared to the UST group (Standard MD = -2.90, CI = -7.51 to 1.72, P = 0.22). A high level of heterogeneity was found amongst the studies (I^2^ = 99%, P <0.00001).

**Figure 4:**
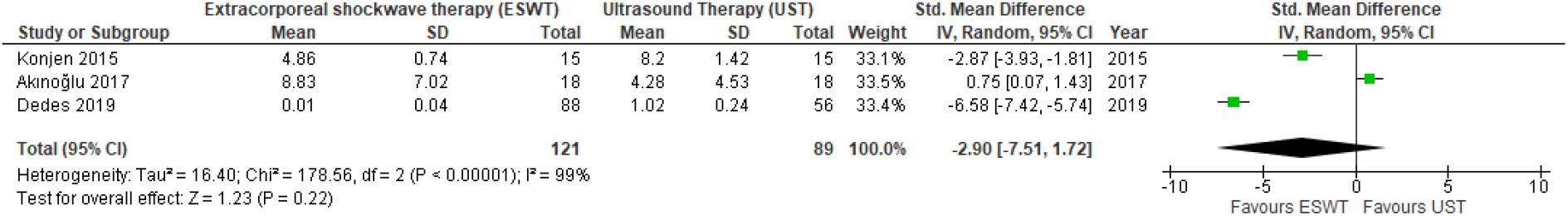
Forest Plot of ESWT vs. UST – Functional impairment. Quantitative analysis showing the standard mean difference in functional impairment reported by Konjen et al. (2015), Akınoğlu et al. (2017) and Dedes et al. (2019). ESWT and UST stand for extracorporeal shockwave therapy and ultrasound therapy, respectively.

#### AOFAS Scale Score

In Figure 5, the AOFAS scale score was reported by Ulusoy et al.^24^ and Akınoğlu et al. (2017) ^25^ enrolling 38 patients. There was no statistically significant difference seen in the standard mean difference analyses, showing a higher AOFAS scale score for the UST group (MD = 35, CI = -1.78 to 8.38, P = 0.20). A low level of heterogeneity was found amongst the studies (I^2^ = 0%, P = 0.40).

**Figure 5:**
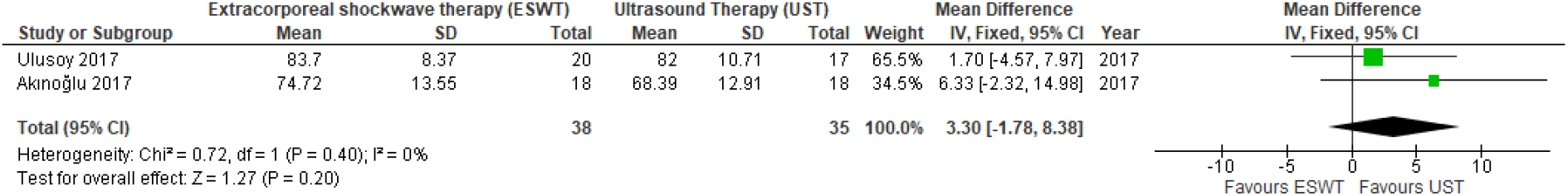
Forest Plot of ESWT vs. UST – AOFAS scale score. Quantitative analysis showing the standard mean difference in UST reported by Ulusoy et al. (2017) and Akınoğlu et al (2017). ESWT and UST stand for extracorporeal shockwave therapy and ultrasound therapy, respectively.

### Secondary outcomes

#### Fascial Thickness

According to Ulusoy et al.^24^, fascial thickness was measured on MRI from coronal and sagittal planes. There was a significant decrease revealed in the thickness of the fascia in both groups after treatment (P < 0.001), but no statistically significant difference was found between the two groups in the reduction of thickness^24^.

#### Primary Efficacy Success Rate

Ulusoy et al.^24^ used the reduction of heel pain as a measurement of primary efficacy rate, which was detected in 65% of the ESWT group and 23.5% of the UST group. In the comparison, ESWT group was found to be more effective than UST group, with a significant difference found between the two groups (P = 0.012) in the success rate^24^.

#### Activity Limitations

Activity limitations were assessed in three studies using different measurements^24,25,27^. Based on Ulusoy et al.^24^ and Dedes et al.^27^, there was a reduction in the activity limitations in both groups, but in comparison, ESWT treatment modality was more effective than UST (P < 0.05). However, Akinoglu et al.^25^ showed that the reduction in the activity limitations was most marked in the UST group compared to ESWT (P < 0.05).

#### Pain Intensity

Pain intensity after treatment was reported to be significantly (P < 0.05) lower for the UST group than the ESWT group in Akinoglu et al.^25^ and Akinoglu et al^26^. Conversely, Dedes et al.^27^ reported significantly improved results in the ESWT group both immediately after the treatment and after 4-week follow-up (P < 0.001). Both Cheing et al.^21^ and Greve et al.^22^ concluded that ESWT is potentially more effective in reducing pain intensity with no significant difference between the two groups.

#### Treatment Satisfaction

Konjen et al.^23^ reported patient satisfaction to be higher in the rESWT group than the UST, with 80% and 33% of patients respectively rating their treatment satisfaction as “very satisfied”.

### Methodological Quality and Risk of Bias Assessment

The Cochrane Collaboration’s Tool was used to assess the quality of the RCTs included in the study (Table 2). For non-randomised studies, the Newcastle-Ottawa scale^29^ was used to assess its quality which offers a star system for analysis (Table 3). Although Cheing et al.^21^ and dedes et al.^27^ showed a low comparability, the study had a high quality for selection and exposure.

**Table 2.** Bia Analysis of the Randomised Trials using the Cochrane Collaboration’s Tool.

| First Author | Bias | Authors' Judgement | Support for Judgement |
| --- | --- | --- | --- |
| Greve et al.<sup>22</sup> | Domain 1 | Low risk | A list of random numbers used |
|  | Domain 2 | High Risk | Allocated based on the withdrawn number |
|  | Domain 3 | Unclear Risk | No blinding was reported |
|  | Domain 4 | Unclear Risk | No information given |
|  | Domain 5 | Low Risk | No missing outcome data |
|  | Domain 6 | Low Risk | All outcome data reported |
|  | Domain 7 | Low Risk | No other bias detected |
| Konjen et al.<sup>23</sup> | Domain 1 | Low Risk | Block randomisation method using computerised random number generator |
|  | Domain 2 | Low Risk | Used sealed opaque envelopes with random assigned numbers. |
|  | Domain 3 | Unclear Risk | No blinding was reported. |
|  | Domain 4 | Unclear Risk | No information given. |
|  | Domain 5 | Low Risk | Noted all participants leaving or not completing the study. |
|  | Domain 6 | Low Risk | All outcome data reported. |
|  | Domain 7 | Low Risk | No other bias detected. |
| Ulusoy et al.<sup>24</sup> | Domain 1 | Low Risk | One author used the stratified block randomization method. |
|  | Domain 2 | High Risk | Allocated patients according to gender and BMI. |
|  | Domain 3 | High Risk | For practical reasons no blinding was performed for the allocated treatment. |
|  | Domain 4 | Unclear Risk | No information given. |
|  | Domain 5 | Low Risk | Noted all participants leaving or not completing the study. |
|  | Domain 6 | Low Risk | All outcome data reported. |
|  | Domain 7 | Low Risk | No other bias detected. |
| **Akinoğlu et al. (2017)** <sup>25</sup> | Domain 1 | Unclear Risk | No information given. |
|  | Domain 2 | Low Risk | Sealed envelope method was used for randomisation. |
|  | Domain 3 | Low Risk | Single-blinded. |
|  | Domain 4 | Unclear Risk | No information given. |
|  | Domain 5 | Low Risk | Noted all participants leaving or not completing the study. |
|  | Domain 6 | Low Risk | All outcome data reported. |
|  | Domain 7 | High Risk | Only female participants were included. |
| **Akinoğlu (2018)** <sup>26</sup> | Domain 1 | Unclear Risk | No information given. |
|  | Domain 2 | Low Risk | Sealed envelope method was used for randomisation. |
|  | Domain 3 | Low Risk | Single-blinded. |
|  | Domain 4 | Unclear Risk | No information given. |
|  | Domain 5 | Low Risk | Noted all participants leaving or not completing the study. |
|  | Domain 6 | Low Risk | All outcome data reported. |
|  | Domain 7 | High Risk | Only female participants were included. |
| Domain 1 = Random sequence generation (selection bias)<br>Domain 2 = Allocation concealment (selection bias)<br>Domain 3 = Blinding of participants and personnel (performance bias)<br>Domain 4 = Blinding of outcome assessment (detection bias)<br>Domain 5 = Incomplete outcome data (attrition bias)<br>Domain 6 = Selective reporting (reporting bias)<br>Domain 7 = Other bias |  |  |  |

**Table 3.**
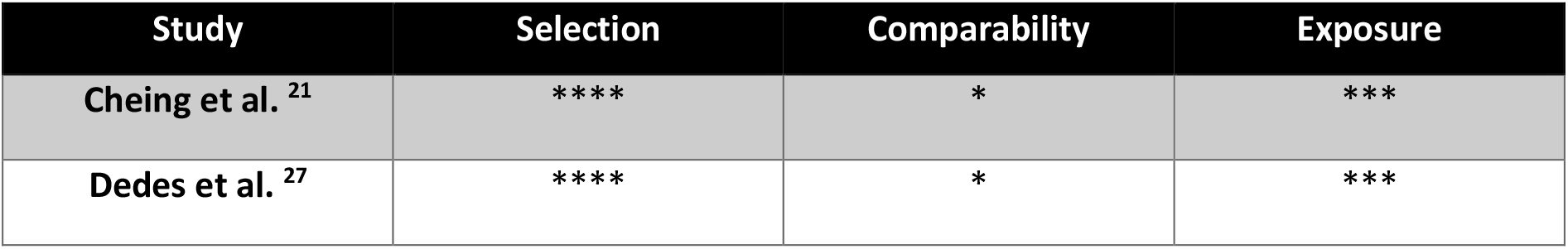
Newcastle-Ottawa scale (NOA) to assess the quality of non-randomised studies.

| Study | Selection | Comparability | Exposure |
| --- | --- | --- | --- |
| **Cheing et al.** <sup>21</sup> | **** | * | *** |
| **Dedes et al.** <sup>27</sup> | **** | * | *** |

## Discussion

ESWT showed a no superior effect when compared with UST in terms of functional impairment, AOFAS scale score and morning pain shown by the results of the analyses. Functional impairment showed no significant (P = 0.22) improvement in the ESWT group compared with the UST group (Figure 4). Similarly, there was no significant (P = 0.20) enhancement in the score of AOFAS scale in the ESWT group (Figure 5). ESWT group showed no significant (P = 0.39) difference in pain during the first steps in the morning when compared with the control group (Figure 2). Conversely, there was a significant (P = 0.005) improvement in the analysis of the pain during activity for the ESWT group (Figure 3). In terms of the between-study heterogeneity, it was low for AOFAS scale score (I^2^ = 0%), moderate for activity pain (I^2^ = 73%) and high for both morning pain (I^2^ = 100%) and functional impairment (I^2^ = 99%) according to the heterogeneity assessment mentioned in the methodological section.

Considering the secondary outcomes, ESWT group showed a significant improvement in primary efficacy success rate (P = 0.012) and activity limitations (P < 0.05) when compared to UST ^24,25,27^. However, in terms of pain intensity after treatment, results varied amongst included studies. A significant (P < 0.05) reduction was reported in favour of UST in two studies^25,26^, while another study revealed a significant (P < 0.001) reduction in pain in ESWT group^27^. Two other studies also concluded that ESWT is potentially more effective in reducing pain intensity with no significant difference found between the two groups ^21, 22^. With respect to reduction of planter fascia thickness, no statistically significant difference was found between the two groups^24^.

There have been multiple studies in the literature about the best choice of treatment for plantar fasciitis. According to the collected data, morning pain and pain during activity were reduced in patients who had both ESWT and UST. The ESWT was even more influential for managing pain during activity compared to UST. Findings from another study^30^ added to the growing number of positive reports that substantiate the effectiveness of ESWT on the treatment of plantar fasciitis by reporting a mean of VAS scores to be decreased from an average of 9.2 to 3.4, at four weeks after treatment. Additionally, a similar RCT found that ESWT had higher pain reduction compared to UST at 3, 6 and 12 weeks after the treatment^31^. The aforementioned studies support the current study’s findings, which can be attributed to the two proposed mechanisms of ESWT, namely the inhibition of painful stimuli resulting in pain relief or the activation of the healing cascade ^15,16^. Considering the data from the available studies, ESWT should be preferred over UST in the management of chronic plantar fasciitis.

A summary of the available evidence was provided in this review using a systematic approach as well as an assessment of the risk of bias of relevant studies and trials^21-24^. Five RCTs and two cohort studies were homogenous based on the included population of interest and design. Therefore, this allows for a non-biased comparison. The combination of these factors makes the conclusions of the current study robust from the best available evidence. Nevertheless, the data of this paper should be studied in terms of inherent limitations. The identification of only seven studies may not be sufficient to make definitive conclusions. Therefore, the findings of the study are exposed to a potential type 2 error.

## Conclusions

Although the evidence is limited with only four studies comparing ESWT and UST, the results of this meta-analysis suggest that ESWT is a superior option in patients with plantar fasciitis. It improves the pain activity and intensity, primary efficacy success rate, activity limitations and patient satisfaction and does not worsen morning pain, functional impairment, AOFAS scale score and the plantar fascial thickness. The authors suggest the requirement of further clinical studies support the current conclusions.

## Data Availability

The datasets generated and analysed during the current study are available from the corresponding author on reasonable request.

## Declarations

### Ethics Approval and Consent to Participate

Not Applicable.

### Consent for Publication

Not Applicable.

### Author Contributions

Z.A. and M.K. contributed equally to the paper as first co-authors for study concept and design as well as contributing in acquisition and analysis of data. E.A. and A.A. interpreted the data and contributed in drafting the manuscript. All authors read and approved the final manuscript. M.K. was also responsible for study supervision.

## Acknowledgements

Not Applicable.

